# Modeling the initial phase of SARS-CoV-2 deposition in the respiratory tract mimicked by the ^11^C radionuclide

**DOI:** 10.1101/2020.07.27.20156836

**Authors:** Heitor Evangelista, Cesar Amaral, Luís Cristóvão Porto, Sergio J. Gonçalves, Eduardo Delfino Sodré, Juliana Nogueira, Angela M.G. dos Santos, Marcio Cataldo

**Affiliations:** LARAMG - Laboratory of Radioecology and Global Changes/DBB/IBRAG. UERJ, Pavilhão Haroldo L. Cunha/DBB/IBRAG. Rua São Francisco Xavier 524. Maracanã, Rio de Janeiro State University, Rio de Janeiro, RJ, 20550-013. Brazil; Piquet Carneiro Polyclinic/Histocompatibility and Cryopreservation Laboratory, Rio de Janeiro State University, Rio de Janeiro State University. Av. Mal. Rondon, 381 - São Francisco Xavier, Rio de Janeiro, RJ, 20950-003. Brazil; Draxos Consultoria e Gestão Ambiental Ltda. Av. das Américas 7938 - Sala 329, Rio de Janeiro, RJ, 22793-081. Brazil

**Keywords:** SARS-CoV-2, COVID-19, respiratory tract, viral load, radionuclides, ICRP-66

## Abstract

The knowledge on the deposition and retention of the viral particle of SARS-CoV-2 in the respiratory tract during the very initial intake from the ambient air is of prime importance to understand the infectious process and COVID-19 initial symptoms. To give some light on that, we propose to use a modified version of a widely tested lung deposition model developed by the ICRP, in the context of the ICRP Publication 66, that provides deposition patterns of microparticles in different lung compartments. In the model, we mimicked the “environmental decay” of the virus, determined by controlled experiments related to normal speeches, by the radionuclide ^11^C that presents comparable decay rates. Our results confirm clinical observations on the high virus retentions observed in the extrathoracic region and the lesser fraction on the alveolar section (in the order of 5), which relevance is a subject to be investigated.

## 1 Introduction

Respiratory infections are relevant clinical conditions due to their diffusion and potentially severe consequences, such as presently observed for the SARS-CoV spread all around the world. Declared as a “global pandemic” by the World Health Organization (WHO) on March 11th, 2020, the new coronavirus disease 2019 (COVID-19) represents one of the greatest public health challenges in recent decades (1). Respiratory infections caused by viruses, mainly due to their high capacity of infection and spread, are a major cause of illness and death from a global perspective. The WHO has included coronaviruses into this group with the global alert caused by the SARS (Severe Acute Respiratory Syndrome) epidemic in 2003, caused by a newly described coronavirus (SARS-CoV). Since then, several other pathogens associated with acute respiratory system disorders have been described, such as more aggressive strains of Influenza, MERS-CoV in the Middle East, and also new types of coronavirus NL63 and KHU1 (2,3). Thus, understanding the modes of transmission of these emerging infectious diseases is a key factor both for the protection of health workers, who deal with infected people, as well as for the public, that sooner or later will be exposed to places where these agents circulate.

Airborne transmission is a key issue for the understanding of SARS-CoV-2 spread out, and it is particularly important for the community of healthcare professionals since they are more exposed to infected patients. However, it is not yet fully clarified the potential of contagion for the general public related to the high agglomeration of people, as observed in various modes of urban transport in large cities, especially in underdeveloped countries. In Italy (Bergamo city), a first study showed that RNA SARS-CoV-2 might be present in association with microparticles in the outdoor environment. Still, the detection of the virus itself was mostly inconclusive (4). Once in the free atmosphere, virus particles are submitted to conditions of natural denaturation or inactivation due to solar radiation, relative humidity, and air temperature (5,6). Such conditions result in dehydration of the viral particles derived from speech, sneezing, or coughing and, as a consequence, becoming an agglutination of viral particles with others suspended organic and inorganic molecules/microparticles in the air. Such interactions lead to changes in its size distribution and consequently effecting its diffusion/dispersion/deposition and residence time in the air. Unlike other stochastic models that determine the deposition fractions in the lung compartments from aerodynamic properties of aerosols, we present here a simulation of a model that considers the full biokinetics of the radionuclide ^11^C mimicking particles containing viruses. To perform that, we used a modified version of a widely tested lung deposition model developed by the International Commission on Radiological Protection (ICRP), in the context of the ICRP Publication 66 (Human Respiratory Tract Model for Radiological Protection-66) from the ICRP Task Group on Internal Dosimetry.

## 2 Methods

During the last decades, several studies have deeply investigated the dynamics of respiratory infections to gain information on effective treatment/prevention of these clinical events (7). Many different models have been developed till then. A respiratory infection model is a system that emulates the complexities observed on the relationship between the infectious agents and the host’s defenses. Several *in vivo* and *in vitro* respiratory models for humans and other vertebrates are available and could be easily reproduced. Mathematical models are also available and were created to numerically describe the principles and evolution of respiratory infections and their diffusion. All of them require specific inputs and have complexities related to the condition they attempt to emulate, therefore presenting both advantages and limitations.

### 2.1 The ICRP Model

The ICRP has developed models for aerosol pulmonary deposition, given the intensive use of natural and artificial radionuclides in the nuclear industry. Such activities range from uranium mining, where workers are exposed to dust containing naturally occurring radioactive aerosols (8), to sectors where there is the handling of wastes from nuclear power plants and related facilities (9,10). Such models depend on the aerosol size distribution of the radioactive aerosols suspended in the air, their chemical form, and its solubility and the corresponding biokinetic processes associated (11). In this context, extensive studies on radioactive aerosols biokinetics lead to successful practical application and have been particularly improved in calculating the internal dose of workers and individuals, especially those living in regions with high levels of natural radioactivity (12).

Herein we use a radionuclide-based model starting with the premise that the virus (SARS-Cov-2) has an “environmental half-life” due to its degradation process in the environment before the deposition in the respiratory tract. From this assumption, we draw a parallel with the half-life of a radionuclide, and thus being able to attribute to the virus an analog “constant of disintegration” related to the outdoor environment. The choice of the radionuclide was based on the study of Stadnytskyi et al. (13), that used a highly sensitive laser light scattering system to track the dynamics in the air of speech droplets generated by carriers of severe acute respiratory syndrome coronavirus 2 (SARS-CoV-2). Droplets were produced during the normal speaking condition. Speech droplets can be suspended in the air for a tenth of minutes and, therefore, can be a potential source of airborne virus transmission in low disturbed places. The settling down (vanishing) time scale of droplets before dehydration and attachment to other existing microparticles in the environment describes a behavior close to the disintegration curve of the artificial radioisotope ^11^C, which half-life is 20.33 minutes, Fig.1a. ^11^C is one of the most useful radionuclides employed for Positron Emission Tomography (PET) radiochemistry because its attachment to a biologically active molecule does not modify the biochemical properties of the inoculated compound (14). From the above, we elected the ^11^C to mimic the virus intake and deposition in the respiratory tract. Another reason relies on the fact that carbon molecules are one of the base constituents of cells and tissues; this allows very precise information on metabolism processes, receptor/enzyme function, and biochemical mechanisms. As a basic complementary input to the model, we used airborne SARS-CoV-2 size distribution data (15) in a stand of theoretical curves or data derived from other virus types as influenza, Fig. 1b. They obtained the size distribution of viral particles from a sequence of speeches in pre-sterilized gelatin filters installed inside a miniature cascade impactor during an air monitoring in the Renmin Hospital of Wuhan University - China, in February-March 2020. In their study, MMAD varied from 5.0 to 5.5 μm. Other studies of droplets produced during speeches with sustained vocalization found modes from 1.8 to 5.5 μm (16).

**Figure 1.**
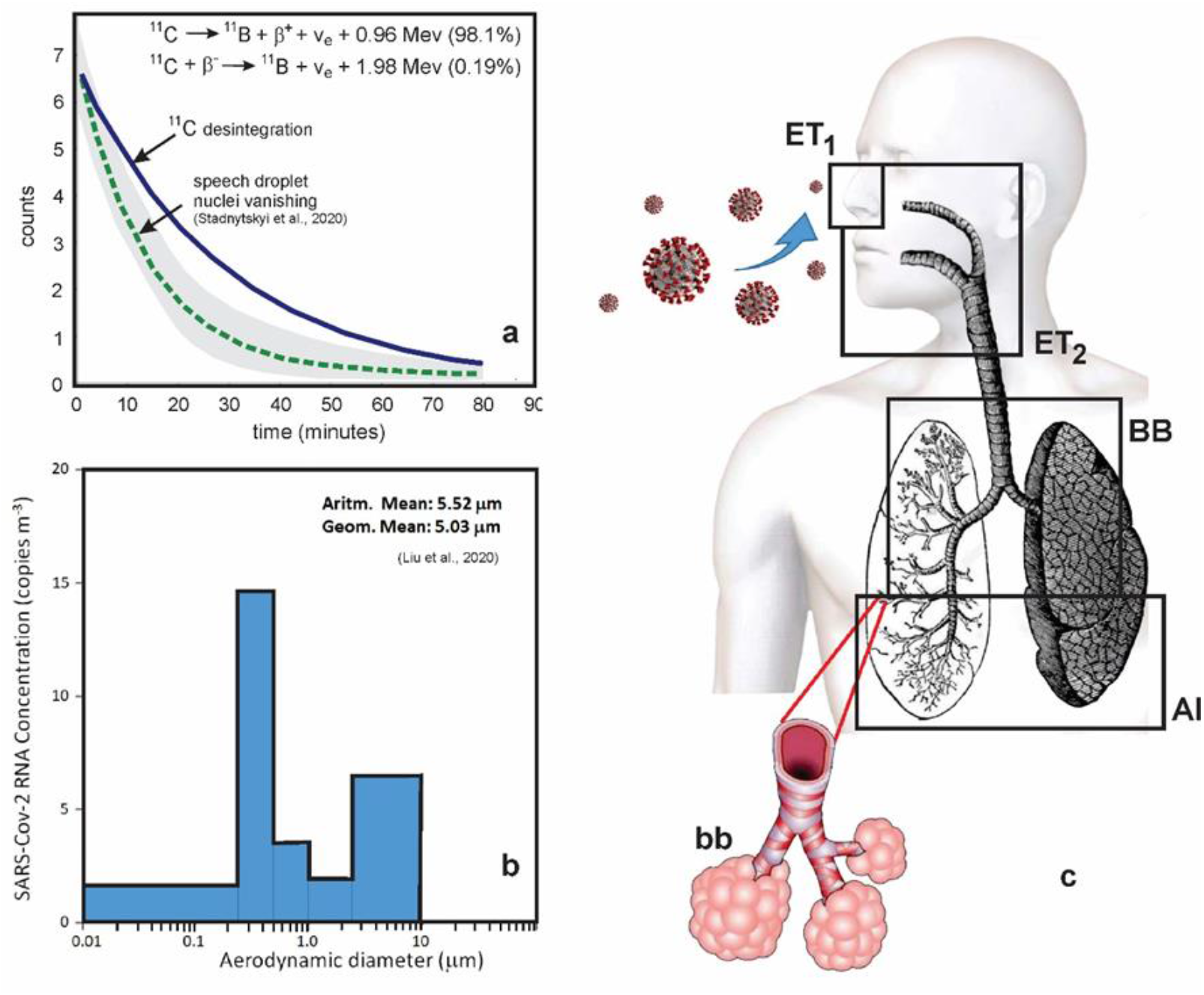
Basic inputs for the lung deposition model: (a) disintegration curve of the radionuclide ^11^C (in bold) and speech droplet nuclei vanishing (dotted line) and variance (gray shaded area); (b) size distribution of SARS-Cov-2 obtained by Liu et al., (2020); (c) respiratory tract compartments investigated by the model: AI (alveolar), bb (bronchioles), BB (bronchi), ET_1_ (extrathoracic region 1 - retention of material deposited in the anterior nose) and ET_2_ (extrathoracic region 2 – retention of material deposited in the posterior nasal passage, larynx, pharynx, and mouth).

An imperative assumption to allow the use of the ICRP model, which is specially designed for radionuclides use, is to attribute a “decay property” to the virus. However, since the virus no longer is submitted to any “decay” after its intake by the human organism, the assumption of mimicking by the ^11^C stops at that stage. The number concentration of active virus particles inoculated in a body remains the same until the physiological responses start, and virus elimination/cell infections and replication takes place. This means that our estimates refer only to the initial deposition of the virus, and therefore the deposition fractions presented here were calculated for the first 1 minute after deposition.

The model used subdivides the respiratory system into two basic compartments: the extrathoracic region (composed of nose and trachea) and the thoracic region (composed of the bronchi to the alveolar sacs). The compartments AI, bb, BB, ET_1_, and ET_2_ represent the alveolar, bronchioles, bronchi, and extrathoracic regions, respectively, Fig. 1c. The lung is considered to have 16 generations where bifurcations occur in the bronchial and bronchioles tree. The BB region comprises the generations from 0 to 8 and the bb from 9 to 15. The rates of mechanical removal of a compartment are expressed in 1/time. Ciliary transport and retention time in the tissues migrating to the pulmonary lymph nodes are also considered in the model, as well as blood absorption from these compartments, causing then a dispute between mechanical removal and blood absorption. The compartments above act as source compartments in the infection of the lung tissues. All the calculations were performed by the AIDE (Activity and Internal Dose Estimates) software (17). The basic inputs of the model are presented in Table 1, where AMAD means “aerosol containing the viral load” median aerodynamic diameter, what implies that 50% of the viral load in the aerosol is associated with particles of aerodynamic diameter greater than the AMAD (used when deposition depends mainly on inertial impaction and sedimentation). At the same time, a compound refers to a material classified according to its rate of absorption from the respiratory tract to body fluids. In this case, deposited Type F materials are those that are readily absorbed into body fluids from the respiratory tract (Fast absorption), and f_1_ is the fractional absorption in the gastrointestinal tract.

**Table 1.** Specifications of model parameters and inputs.

| AIDE model parameters | AIDE model inputs |
| --- | --- |
| **Model** | C, Isotope: C-11 |
| **Data source** | Carbon according to ICRP-67* |
| **Subject** | Reference Worker (breathing rate of $1.2 \text{ m}^3 \text{ h}^{-1}$) |
| **Intake type** | Inhalation, Single |
| **Initial or Daily Activity** | $1.000\text{E}+00 \text{ Bq}$ |
| **Inhalation type** | Respiratory Tract model: ICRP-66 |
| **Compound** | Type F |
| **GI Tract Absorption Factor** | $f_I = 1.000\text{E}+00$ |
| **AMAD ($\mu\text{m}$)** | $5.000\text{E}+00^{**}$ |

\* ICRP, 1993. Age-dependent Doses to Members of the Public from Intake of Radionuclides – Part 2 Ingestion Dose Coefficients. ICRP Publication 67. Ann. ICRP 23 (3-4)
\*\* Based on the geometric mean of Fig.1b.

### 2.2 SARS-Cov-2 Field Data

Because COVID-19 is highly associated through the respiratory airways, we also present data of nasopharyngeal swabs (1490) and bronco-tracheal aspirate (4) COVID-19 RT-PCR tests performed at Pedro Ernesto University Hospital (HUPE), one of the specialized units for COVID-19 treatment in the Rio de Janeiro City/Brazil. We used the data to observe the evolution of the pandemics moments just before and after the mandatory use of personal safety equipment (masks) has been implemented by the local government. The database for patients with COVID-19 started on March 29^th^ (13^th^ epidemiological week) up to July 12^th^ (28^th^ epidemiological week) when 939 patients were hospitalized and a total of 1,494 individuals were tested on this hospital. Among the realized tests, all the four bronco-tracheal aspirates resulted in negative for SARS-CoV-2, the remaining test results were presented in Table 2.

**Table 2.** Number of detected new and total tests in Pedro Ernesto Univesity Hospital from Epidemiological week 13 to 28.

| Epidemiological Week | Date | Hospitalizations |  |  | Total |  |  |
| --- | --- | --- | --- | --- | --- | --- | --- |
|  |  | Tests | Detected | (%) | Tests | Detected | (%) |
| 13 | 29-mar | 11 | 2 | 18.2 | 11 | 2 | 18.2 |
| 14 | 5-abr | 22 | 7 | 31.8 | 24 | 8 | 33.3 |
| 15 | 12-abr | 40 | 16 | 40.0 | 48 | 19 | 39.6 |
| 16 | 19-abr | 46 | 40 | 87.0 | 58 | 50 | 86.2 |
| 17 | 26-abr | 42 | 36 | 85.7 | 47 | 40 | 85.1 |
| 18 | 3-mai | 86 | 52 | 60.5 | 112 | 63 | 56.3 |
| 19 | 10-mai | 93 | 55 | 59.1 | 142 | 74 | 52.1 |
| 20 | 17-mai | 56 | 31 | 55.4 | 108 | 57 | 52.8 |
| 21 | 24-mai | 53 | 27 | 50.9 | 91 | 35 | 38.5 |
| 22 | 31-mai | 106 | 46 | 43.4 | 179 | 74 | 41.3 |
| 23 | 7-jun | 74 | 21 | 28.4 | 117 | 29 | 24.8 |
| 24 | 14-jun | 72 | 19 | 26.4 | 119 | 30 | 25.2 |
| 25 | 21-jun | 54 | 8 | 14.8 | 117 | 17 | 14.5 |
| 26 | 28-jun | 59 | 10 | 16.9 | 126 | 26 | 20.6 |
| 27 | 5-jul | 55 | 4 | 7.3 | 91 | 7 | 7.7 |
| 28 | 12-jul | 70 | 5 | 7.1 | 104 | 9 | 8.7 |

## 3 Results

Our results point to a far more relevant deposition fraction at the extrathoracic compartments of ET1 accounting to 47.4 and ET2 to 48.78 for the virus intake after 1 minute. In contrast, the sum for the bronchial and bronchioles compartments corresponded to 3.75 (Fig. 2). Data generated by the model is provided in the Supplementary Material.

**Figure 2.**
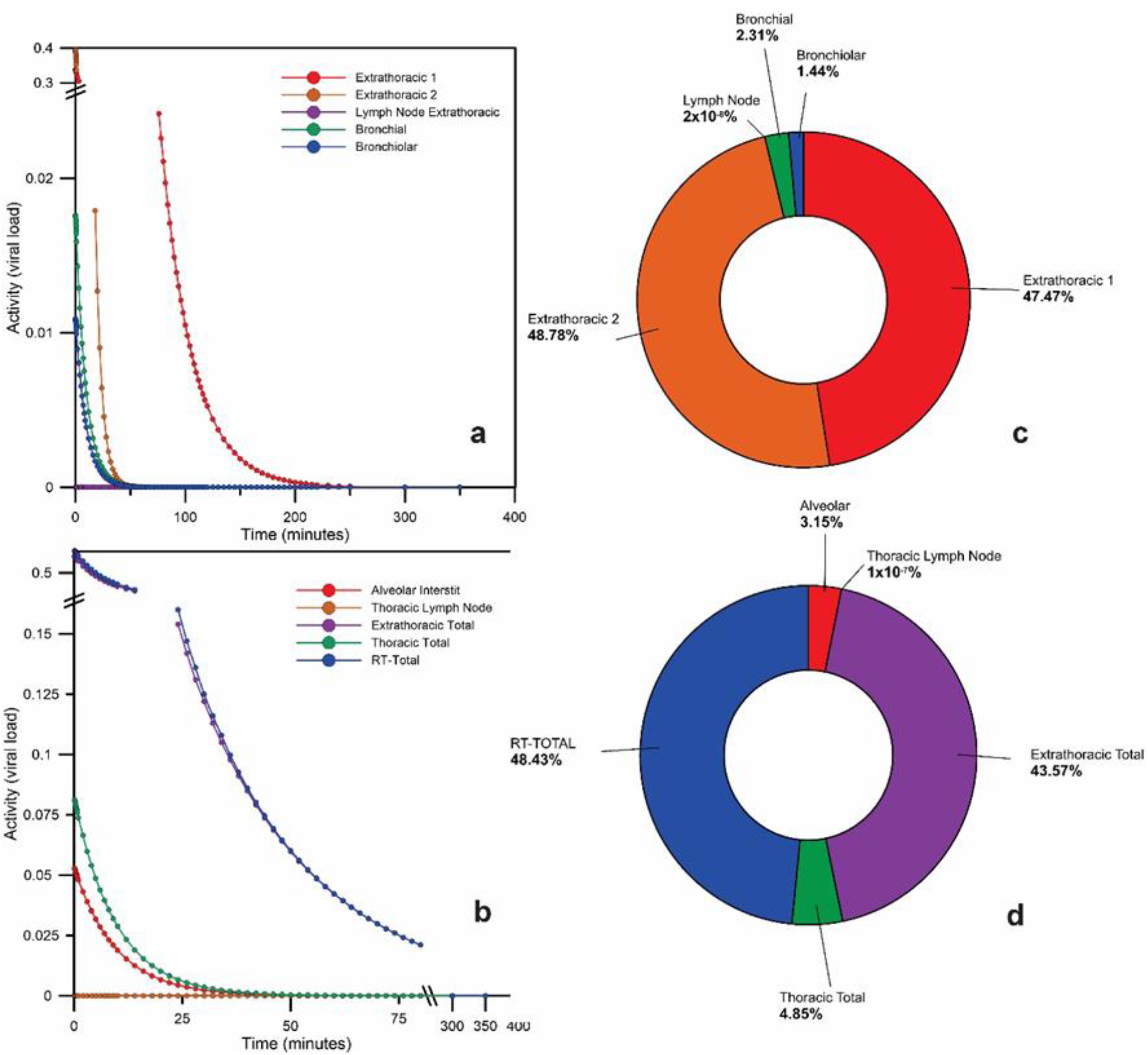
Time-dependent particle transport from each respiratory tract region in the compartments of the model for SARS-Cov-2 according to the ICRP model as time response curves (a and b); and fractions of the initial intake of the virus by ET_1_ pathway after 1 minute of exposure (c and d).

## 4 Discussions

Our results point to a far more relevant deposition fraction at the extrathoracic compartments of ET_1_ accounting for 47.4% and ET_2_ to 48.78% for the virus intake after 1 minute of exposure. In contrast, the sum for the bronchial and bronchioles compartments corresponded to 3.75% (Fig. 2). For a comparison with a Stochastic Lung Deposition Model, as proposed by Madas et al. (18), they found 61.8% of the total inhaled mass fraction to be at the upper airways, ∼8.5% for the acinar airways and ∼5.5% for the bronchial compartment corresponding to a single inhalation. The differences between the two models can be attributed to the fact that Madas et al. (18) have used a mass size distribution of particles from influenza with modal values between 2-3 µm, derived from coughing of patients while we used a SARS-Cov-2 data in conditions of speeches with AMAD ∼ 5 µm. Our results stress the impact of the upper airways in the initial airborne virus retention in the respiratory tract since the total extrathoracic compartment may retain more than 96% of the virus load. Both models point to at least ∼4% of contribution to the most inner parts of the lung (bronchial and bronchioles). In the SARS-Cov-2 size distribution we have used, it is evident an existing viral load in the ultrafine particle size range (< 0.1 mm diameter). Their behavior differs from other groups of particles as the fine and coarse modes, since the virus attached to ultrafine particles may be deposit in the inner lung compartments by diffusion mechanisms. From controlled experiments, it is known that ultrafine particles peak deposition occurs in lung regions that encompass the transition zone between the conducting airways and the alveolar region (19). Therefore, since our model predicts a small fraction of particles containing virus reaching directly to the alveolar region its importance in the development of the disease should be considered since SARS-Cov-2 may bind to the cells at that compartment through ACE2 (Angiotensin-Converting Enzyme 2) receptors that are the host cell receptor responsible for mediating infection by SARS-CoV-2 (20). ACE2 plays an important role in the lungs protecting it from acute respiratory distress syndrome (ARDS) by breaking down Angiotensin II, which has inflammatory effects (21,22). Binding of the SARS-CoV-2 to ACE2 inhibits it and thus weakens its protective action on the organ (23).

The pathogenesis and virus transmission pathways are still being investigated and the already published results are, in fact, still under intense debate. That is the case for the airborne transmission of the virus. The analysis of the temporal dynamics of the SARS-Cov-2 infection suggests that after 2-3 days of the first symptoms the viral shedding starts, therefore favoring the pre-symptomatic/asymptomatic transmission of the virus (24). In this sense, understanding the first phases of the virus infection is paramount. Our results were derived from normal speech conditions as viral source-term and indoor conditions. In this specific case, we observed extrathoracic percentage infection levels in good agreement with clinical observations of patients that initially presented mild COVID-19 symptoms evolving to a more deteriorated health board (25). Although COVID-19 manifestation related to a minimum infectious dose, like that expected to the alveolar compartment, is so far not clear, and considering that: (a) none masks have 100% retention efficiency; (b) SARS-CoV-2 viral particles are found in aerodynamic diameters shorter than 0.1 μm in surveys; (c) model predicts around 5% direct virus insertion at the alveolar compartment, these facts when combined may explain why a fraction of the population even using masks get contaminated with “no apparent reason”. Respiratory tract models developed by the ICRP Task Groups have been largely used for safety and protection in nuclear activities in several countries reaching excellent performance and being validated by internal measurements. Their use for non-radioactive aerosols of biological and mineral origin and pollutants is an emerging topic and a potential to be explored.

Since ET1 and ET2 are key-compartments in the initial phase of SARS-Cov-2 deposition, we investigated the impact of the use of individual protection such as masks and face shields on the epidemiological data related to the COVID-19. Table 2 shows a survey developed by our working group from nasopharyngeal swabs and bronco-tracheal aspirate tests for COVID-19 by RT-PCR performed at Pedro Ernesto University Hospital (HUPE), in Rio de Janeiro, Brazil. The city of Rio de Janeiro in no time had adopted a full lockdown strategy, but just recommendations of social distance and personal care such as hands washing and alcohol use for personal and items/objects disinfection. However, COVID-19 data showed continuous prevalence forcing the local authorities to carry out a mandatory use of masks on April 23^rd^. Our results clearly suggest that the fraction of both hospitalized and total tested patients with SARS-CoV-2 detected from the RT-PCR tests exhibited a significant decrease when we observed an immediate drop in percentage from 85% to 60% (Figure 3). We do believe that the analyzed sample is representative of the city population since the Pedro Ernesto University Hospital (HUPE) receives patients from several health units spread all over the urban domain. We should also note that due to socio-economic reasons and also difficulties in the acquisition of high-quality safety equipment, most of the population made use of simple, cheap, and home-made masks and face shields. However, despite these circumstances, effective response against the pandemic was achieved.

**Figure 3.**
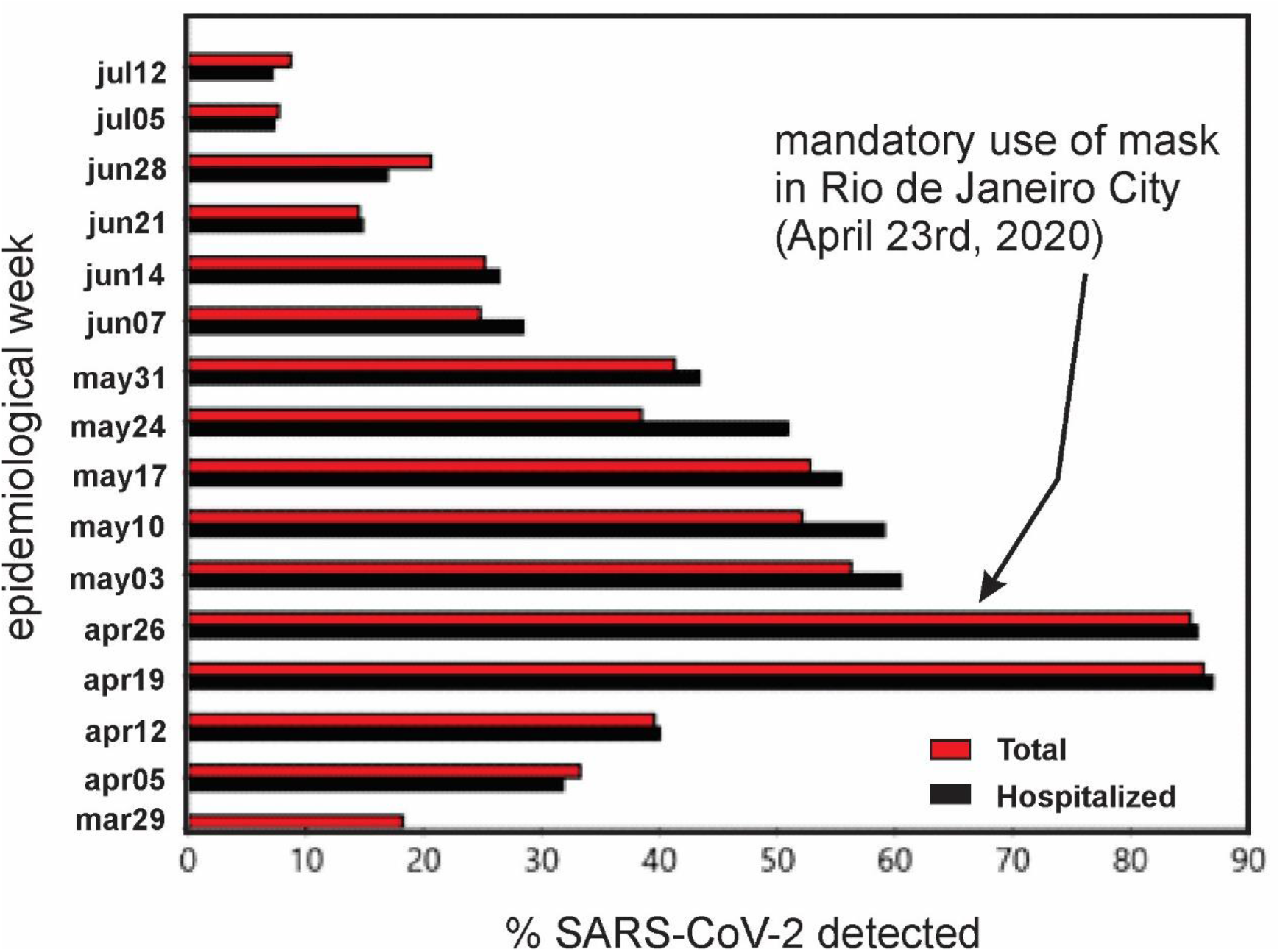
SARS-CoV-2 detection from nasopharynx COVID-19 RT-PCR tests performed at Pedro Ernesto University Hospital (HUPE) in Rio de Janeiro/Brazil.

In summary, respiratory tract models developed by the IAEA Group tasks have been largely used for safety and protection in nuclear activities in several countries, reaching excellent performance and being validated by *in locus* measurements. Their use for non-radioactive aerosols and microparticles of biological and mineral origin and pollutants is an emerging topic and a potential to be explored.

## Data Availability

All data used to generate the results presented herein are provided in the supplementary material.

## Conflict of Interest

The authors declare that the research was conducted in the absence of any commercial or financial frameworks that could be construed as a potential conflict of interest.

## Author Contributions

All authors contributed equally to the development and text of the manuscript. Heitor Evangelista (Project coordinator, creator of the project), Luís Cristóvão Porto (medical survey), Sergio Gonçalves Junior (participation in the text content and discussions), Eduardo Delfino Sodré (participation in the text content and discussions), Juliana Nogueira (numeric model analysis), Cesar Amaral (participation in the text content and discussions), Marcio Cataldo Gomes da Silva (participation in the text content and discussions).

## Funding

The authors received no funding for this research.

## Acknowledgments

We greatly thank Dr. Luiz Bertelli for the availability of AIDE (Activity and Internal Dose Estimates) software and important comments on the numeric model and Roberta Priori for draw designs.

## Notes

### Competing Interest Statement

The authors have declared no competing interest.

### Author Declarations

The use of COVID-19 data was approved by the Ethics in Research Committee of the Pedro Ernesto University Hospital under the project entitled Epidemiological, laboratorial, and clinical profile of the COVID-19 pandemic of patients treated at Rio de Janeiro State University (UERJ) (CAAE: 30135320.0.0000.5259). All COVID-19 typing data were de-identified prior to the analysis and available for the authors uniquely as totals, as presented on Table 2.

## References

1. Santarpia JL, Rivera DN, Herrera V, Morwitzer MJ, Creager H, Santarpia GW, et al. Aerosol and Surface Transmission Potential of SARS-CoV-2. MedRxiv. 2020;

2. Van der Hoek L, Pyrc K, Jebbink MF, Vermeulen-Oost W, Berkhout RJM, Wolthers KC, et al. Identification of a new human coronavirus.Nat Med [Internet]. 2004 Apr 21;10(4):368–73. Available from: http://www.nature.com/articles/nm1024

3. Woo PCY, Lau SKP, Chu C, Chan K, Tsoi H, Huang Y, et al. Characterization and Complete Genome Sequence of a Novel Coronavirus, Coronavirus HKU1, from Patients with Pneumonia. J Virol [Internet]. 2005 Jan 15;79(2):884–95. Available from: https://jvi.asm.org/content/79/2/884

4. Setti L, Passarini F, De Gennaro G, Barbieri P, Perrone MG, Borelli M, et al. SARS-Cov-2RNA found on particulate matter of Bergamo in Northern Italy: First evidence. Environ Res [Internet]. 2020 Sep;188:109754. Available from: https://linkinghub.elsevier.com/retrieve/pii/S0013935120306472

5. Chan KH, Peiris JSM, Lam SY, Poon LLM, Yuen KY, Seto WH. The effects of temperature and relative humidity on the viability of the SARS coronavirus. Adv Virol. 2011;

6. Ratnesar-Shumate S, Williams G, Green B, Krause M, Holland B, Wood S, et al. Simulated Sunlight Rapidly Inactivates SARS-CoV-2 on Surfaces. J Infect Dis [Internet]. 2020 Jun 29;222(2):214–22. Available from: https://academic.oup.com/jid/article/222/2/214/5841129

7. Saturni S, Contoli M, Spanevello A, Papi A. Models of Respiratory Infections: Virus-Induced Asthma Exacerbations and Beyond. Allergy Asthma Immunol Res [Internet]. 2015;7(6):525. Available from: https://e-aair.org/DOIx.php?id=10.4168/aair.2015.7.6.525

8. Otahal P, Burian I. The airborne natural radioactivity in the uranium mine Rozna I. Radiat Prot Dosimetry [Internet]. 2011 May 1;145(2–3):150–4. Available from: https://academic.oup.com/rpd/article-lookup/doi/10.1093/rpd/ncr046

9. Breustedt B, Giussani A, Noßke D. Internal dose assessments – Concepts, models and uncertainties. Radiat Meas [Internet]. 2018 Aug;115:49–54. Available from: https://linkinghub.elsevier.com/retrieve/pii/S1350448718300143

10. Oberdorster G. Airborne cadmium and carcinogenesis of the respiratory tract. Scandinavian Journal of Work, Environment and Health. 1986.

11. Cuddihy RG, McClellan RO, Griffith WC. Variability in target organ deposition among individuals exposed to toxic substances. Toxicol Appl Pharmacol. 1979;

12. Henricsson F, Persson BRR. Polonium-210 in the bio-sphere: Bio-kinetics and biological effects. In: Radionuclides: Sources, Properties and Hazards. 2012.

13. Stadnytskyi V, Bax CE, Bax A, Anfinrud P. The airborne lifetime of small speech droplets and their potential importance in SARS-CoV-2 transmission. Proc Natl Acad Sci [Internet]. 2020 Jun 2;117(22):11875–7. Available from: http://www.pnas.org/lookup/doi/10.1073/pnas.2006874117

14. Yang J, Feng Y, Zhan H, Liu J, Yang F, Zhang K, et al. Characterization of a Pyrethroid-Degrading Pseudomonas fulva Strain P31 and Biochemical Degradation Pathway of D-Phenothrin. Front Microbiol [Internet]. 2018 May 16;9. Available from: http://journal.frontiersin.org/article/10.3389/fmicb.2018.01003/full

15. Liu Y, Ning Z, Chen Y, Guo M, Liu Y, Gali NK, et al. Aerodynamic analysis of SARS-CoV-2 in two Wuhan hospitals. Nature [Internet]. 2020 Jun 27;582(7813):557–60. Available from: http://www.nature.com/articles/s41586-020-2271-3

16. Morawska L, Johnson GR, Ristovski ZD, Hargreaves M, Mengersen K, Corbett S, et al. Size distribution and sites of origin of droplets expelled from the human respiratory tract during expiratory activities. J Aerosol Sci [Internet]. 2009 Mar;40(3):256–69. Available from: https://linkinghub.elsevier.com/retrieve/pii/S0021850208002036

17. Bertelli L, Melo DR, Lipsztein J, Cruz-Suarez R. AIDE: internal dosimetry software. Radiat Prot Dosimetry [Internet]. 2008 Feb 29;130(3):358–67. Available from: https://academic.oup.com/rpd/article-lookup/doi/10.1093/rpd/ncn059

18. Madas BG, Füri P, Farkas Á, Nagy A, Czitrovszky A, Balásházy I, et al. No Title. MedRxiv. 2020;

19. Kim CS, Jaques PA. Respiratory dose of inhaled ultrafine particles in healthy adults. Brown LM, Collings N, Harrison RM, Maynard AD, Maynard RL, editors. Philos Trans R Soc London Ser A Math Phys Eng Sci [Internet]. 2000 Oct 15;358(1775):2693–705. Available from: https://royalsocietypublishing.org/doi/10.1098/rsta.2000.0678

20. Zhang H, Penninger JM, Li Y, Zhong N, Slutsky AS. Angiotensin-converting enzyme 2 (ACE2) as a SARS-CoV-2 receptor: molecular mechanisms and potential therapeutic target. Intensive Care Med [Internet]. 2020 Apr 3;46(4):586–90. Available from: http://link.springer.com/10.1007/s00134-020-05985-9

21. Imai Y, Kuba K, Rao S, Huan Y, Guo F, Guan B, et al. Angiotensin-converting enzyme 2 protects from severe acute lung failure. Nature [Internet]. 2005 Jul;436(7047):112–6. Available from: http://www.nature.com/articles/nature03712

22. Jia H. Pulmonary Angiotensin-Converting Enzyme 2 (ACE2) and Inflammatory Lung Disease. Shock [Internet]. 2016 Sep;46(3):239–48. Available from: https://journals.lww.com/00024382-201609000-00003

23. Kuba K, Imai Y, Rao S, Gao H, Guo F, Guan B, et al. A crucial role of angiotensin converting enzyme 2 (ACE2) in SARS coronavirus–induced lung injury. Nat Med [Internet]. 2005 Aug 10;11(8):875–9. Available from: http://www.nature.com/articles/nm1267

24. He D, Zhao S, Lin Q, Zhuang Z, Cao P, Wang MH, et al. The relative transmissibility of asymptomatic COVID-19 infections among close contacts. Int J Infect Dis [Internet]. 2020 May;94:145–7. Available from: https://linkinghub.elsevier.com/retrieve/pii/S1201971220302502

25. Wu C, Chen X, Cai Y, Xia J, Zhou X, Xu S, et al. Risk Factors Associated With Acute Respiratory Distress Syndrome and Death in Patients With Coronavirus Disease 2019 Pneumonia in Wuhan, China. JAMA Intern Med [Internet]. 2020 Mar 13; Available from: https://jamanetwork.com/journals/jamainternalmedicine/fullarticle/2763184

